# Nigrostriatal Dopaminergic Neurotransmission and Resilience to Peripheral Systemic Risk Factors for Gait Slowing upon Transition to Uneven Surfaces in Older Adult

**DOI:** 10.1101/2025.01.14.25320551

**Authors:** Lana M Chahine, Andrea Rosso, Ian Troidl, Mary Ganguli, Anne Newman, Steven Cummings, Stephanie Studenski, Brian Lopresti, Sarah Royse, Theodore Huppert, Mark Redfern, Patrick J Sparto, Nico Bohnen, Caterina Rosano

## Abstract

Identifying mechanisms that compensate for slow gait speed in older adults is crucial. Dopaminergic neurotransmission curbs deleterious associations of cerebrovascular disease with gait, but whether it compensates for peripheral systemic risk factors (PSRF) for gait slowing has not been studied. In this cross-sectional study of community-dwelling older adults, we examined the relationship between nigrostriatal dopaminergic terminal integrity and gait speed in individuals with and without ≥1 PSRF for gait slowing: obesity, joint pain, or reduced muscle strength. The primary outcome was percent difference in gait speed (%DGS) on transition from even to uneven surface. Participants underwent dopaminergic imaging with dihydrotetrabenazine [^11^C]DTBZ positron emission tomography. Among 197 individuals, (mean (SD) age 74.92 (4.53) years; 61.93% female; 90.86% White), 130 (65.99%) had ≥1 PSRF. Relationship between posterior putamen [^11^C]DTBZ binding and %DGS was modified by PSRF; in those with ≥1 PSRF (but not in those with no PSRF), posterior putamen [^11^C]DTBZ binding was associated with %DGS (β=0.198, p=0.03) independent of potential confounders. This cross-sectional study indicates that striatal dopaminergic neurotransmission may offer resilience to effects of PSRF on gait slowing.

## 1. Introduction

With increasing age, gait becomes slower and less automated, requiring more attention (Rosso et al., 2013; Sorond et al., 2015). These changes in gait have consequences such as increased risk of falls, disability(Perera et al., 2016), and premature mortality (Studenski et al., 2011). The cause of gait slowing in older adults is multifactorial. It can be due to peripheral systemic factors including joint pain, obesity, and reduced muscle strength, among others (Alexander, 1996; Bindawas, 2016; Figgins et al., 2021a; Figgins et al., 2021b; Rosso et al., 2015)). In addition, central nervous system factors, including cerebral small vessel disease, and smaller dorsolateral and striatal gray matter volumes, contribute to gait slowing in older adults without diagnosed neurologic disorders (Sharma et al., 2023; Sorond et al., 2015).

Some older adults maintain mobility and do not exhibit substantial gait slowing despite having one or more risk factors for it. Indeed, we have demonstrated that ∼20% of older adults maintain fast walking speed even in the presence of cerebral small vessel disease and peripheral systemic risk factors (PSRF). These individuals may be demonstrating resilience: the maintenance of normal walking speed despite risk factors for slower gait (Rosso et al., 2017). Like gait slowing, resilience is likely also multifactorial. Hypothetically, resilience could result from either normal or supra-normal function in some components that affect gait, which compensate for abnormal function in other components. We posit that dopaminergic neurotransmission is a candidate pathway that confers resilience. The nigrostriatal dopaminergic system plays a critical role in motor control; nigrostriatal dopaminergic neurotransmission regulates the automated execution of overlearned motor tasks via its connections with sensorimotor cortical and subcortical areas (Gepshtein et al., 2014) and dopaminergic neurotransmission positively predicts walking speed (Cham et al., 2008).

We previously found that the genotype in a dopamine-metabolism gene that is associated with higher synaptic dopamine levels (Met/Met catechol-o-methyltransferase) attenuates the negative effects of age-related cerebral small vessel disease on gait speed (Rosso et al., 2018). These findings are consistent with post-mortem evidence that a combination of nigral dopamine neurons loss and cerebral small vessel disease best predict walking impairment in older adults (Buchman et al., 2011; Buchman et al., 2012; Ross et al., 2004). Less is known about the contributions of dopaminergic neurotransmission to compensation for deleterious effects of PSRF on gait slowing. Importantly, dopaminergic neurotransmission is potentially modifiable pharmacologically, thereby offering novel approaches to treat non-resilient older adults in a targeted fashion.

Our objectives were to examine among community dwelling older adults with and without PSRF for gait slowing the relationship between nigrostriatal dopaminergic terminal integrity and change in gait speed (%DGS) when walking on an uneven vs even surface. We selected %DGS as our primary outcome as this complex task can reflect changes in walking speed resulting from real-world perturbations (Menant et al., 2009; Menz et al., 2003; Zukowski et al., 2024), and may be more sensitive to the effects of dopamine compared to other tasks (Tian et al., 2017). We used [^11^C]-a-(+)-[^11^C]dihydrotetrabenazine ([^11^C]DTBZ), a positron emission tomography (PET) radioligand that targets the type-2 vesicular monoamine transporter (VMAT2), as a measure of nigrostriatal dopaminergic terminal integrity (Bohnen et al., 2006; Gupta et al., 2021; Hsiao et al., 2014). Our hypothesis was that higher [^11^C]DTBZ PET binding would be associated with less decline in %DGS, and that this relationship would be of greater magnitude among those with PSRF for gait slowing.

## 2. Methods

### 2.1. Study Sample

This was a cross-sectional study of community dwelling older adults living in Southwestern Pennsylvania. The study sample was recruited from two ongoing population-based prospective longitudinal studies: the Monongahela-Youghiogheny Healthy Aging Team study (MYHAT) and Study of Muscle, Mobility and Aging (SOMMA). Sample identification, recruitment, and inclusion/exclusion criteria have previously been described.

Briefly, MYHAT (Ganguli et al., 2009) is a population-based cohort study of older adults that identified potential participants via age-stratified random sampling from voter registration lists. MYHAT recruited individuals aged 65 and older who reside in selected small towns of southwest PA and were not living in a nursing home. Exclusion criteria are illness or vision/hearing impairment severe enough to preclude study participation, moderate or severe cognitive impairment defined by age-education-corrected scores <21 on the Mini-Mental State Examination(Folstein et al., 1975) or inability to provide informed consent.

SOMMA(Cummings et al., 2023) recruited age-eligible individuals identified via mailing lists, voter registration, or in university-sponsored registries. Oversampling of zip codes with greater racial and ethnic diversity was performed. SOMMA recruited individuals aged 70 and older living in Southwest PA who were able to walk 400 meters at a usual pace. Exclusion criteria were anticoagulation therapy, contraindications to MRI or muscle biopsy, inability to walk 0.25 miles or climb a flight of stairs, active cancer or with an advanced chronic disease such as heart failure, renal failure on dialysis, Parkinson’s disease, or dementia.

All individuals enrolled in MYHAT without self-reported diagnosis of PD were potentially eligible. Individuals in SOMMA who had successfully completed muscle biopsy within the prior 12 months were potentially eligible. For this analysis, we included only cognitively normal participants without diagnosed neurologic disorder; therefore, participants who self-reported stroke or who were classified as having mild cognitive impairment (MCI) or dementia were excluded, as were those with missing primary outcome or predictor in this analysis (see below).

### 2.2. Assessments

The following assessments were administered; parent study-specific methods are detailed below where applicable. All assessments were obtained within 1 year of imaging.

-Demographics: age, race, ethnicity, sex at birth, and education as reported by the participant

-Peripheral Systemic Risk Factors (PSRF): (1) Joint pain for at least 1 month in the preceding year in the feet, toes, ankles, knees or hips was ascertained via self-report by participants. A participant was considered to have lower extremity joint pain if they responded yes to pain in any of these body regions. (2) Body mass index: weight (kg) and height (meters) were measured. Obesity was defined as body mass index ≥30 (3) Grip strength, a measure of muscle strength, was assessed in kilograms with Jamar dynamometer. Participants were asked to rest their arm on the table, bend their elbow, and squeeze the dynamometer at maximal effort. Two trials for each arm 15 seconds apart were performed. For each participant the maximal value of right or left grip strength was determined and low grip strength was defined as being in the lowest sex-specific quartile of the sample from each parent study separately.

-Gait assessment: Time to complete walking of a 15-meter surface was used to compute walking speed in meters per second (m/s) under 2 conditions: (1) usual walking on an even surface. This is a clinically relevant measure(Hardy et al., 2007; Perera et al., 2006; Perera et al., 2016; Studenski et al., 2011) that is sensitive to age-related striatal degeneration, and reliable; (2) usual walking on uneven surface: walking on a surface where 1.5 cm high wood prisms arranged randomly at a density of 26 pieces/m^2^ were placed underneath carpeting(Thies et al., 2005). Percent difference in gait speed (%DGS) when on uneven vs even surface was calculated as (speed on uneven – speed on even) / speed on even. A negative percent change indicates the individual slowed down.

-Brain imaging: 3 Tesla magnetic resonance (MR) and [^11^C]DTBZ PET were simultaneously acquired on a whole-body human scanner that integrates a 3T MR and PET scanner (Siemens Biograph mMR, Siemens Medical Solutions USA, Malvern, PA).

*PET protocol*: [^11^C]DTBZ was synthesized as previously described (Kilbourn et al., 1995) with high molar activity (59.9±26.3 MBq/nmol at time-of-injection, 3.81±2.42 μg injected mass). [^11^C]DTBZ (573±111 MBq) was injected intravenously as a slow bolus (20-30 sec) and list-mode acquisition of PET emission data commenced at the start of radiotracer injection and continued for 60 minutes. 3T MR images included T1-weighted magnetization-prepared rapid gradient echo (MPRAGE) and Dixon sequences (Dixon, 1984). PET emission data were reconstructed using filtered back projection (FBP) with fourier rebinning into a dynamic series of 20 frames increasing in duration from 15 sec to 300 sec. Attenuation correction of PET emission data was performed analytically using previously described model-based methods (Izquierdo-Garcia et al., 2014; Koesters et al., 2016). Indices of [^11^C]DTBZ specific binding were derived using the non-invasive Logan graphical analysis method (Logan et al., 1996) that estimates the radiotracer distribution volume ratio (DVR), which is defined as the equilibrium ratio of the total distribution volume (V_T_) in target tissues to the V_T_ of a reference region devoid of VMAT2 (occipital cortex). The [^11^C]DTBZ imaging protocol and analysis method are simple to implement and well tolerated by participants with excellent reproducibility and reliability for estimating [^11^C]DTBZ specific binding(Chan et al., 1999; Sossi et al., 2000). For the present study, we report the [^11^C]DTBZ binding potential relative to non-displaceable binding, BP_ND_ (Innis et al., 2007), which is computed from the DVR using the simple relationship BP_ND_ = DVR-1.

*Striatal subregions examined*: volumes of interest (VOIs) for sampling of PET image data were defined on the MPRAGE MR series using an automated atlas-based method (CIC Atlas, Clinical Imaging Center, Imperial College, London, UK) (Tziortzi et al., 2011) that delineates over 100 cortical and subcortical regions in MR native space. These regions include the functional subdivisions of the striatum: the associative (anterior putamen, PreDorsal Caudate, PostDorsal Caudate), limbic (antero-ventral striatum) and sensorimotor (posterior putamen).(Martinez et al., 2003).

*Cortical volume and white matter hyperintensity quantitation*: MRI volumes were distortion-corrected, registered, and segmented as described in (Glasser et al., 2013) using a combination of FSL and FreeSurfer analysis programs. Total and subcortical gray matter volumes were obtained via FreeSurfer(Dale et al., 1999). Regions were labeled with reference to the Desikan atlas(Desikan et al., 2006). Gray matter atrophy was calculated as total gray matter volume/intracranial volume and subcortical gray matter atrophy as subcortical gray matter volume/intracranial volume. White matter hyperintensity on MRI, a measure of cerebral small vessel disease, was quantitated via automated segmentation methods as previously described(Schmidt and Wink, 2017).

-Cognition: participants were classified as normal cognition, MCI or dementia. In MYHAT, this determination was made based on Clinical Dementia Rating (CDR) scale score of >0. In SOMMA, cognition was assessed with the Montreal Cognitive Assessment (MoCA). Participants were classified as normal, MCI, or dementia based on their MoCA score while accounting for age, sex, race, education level(Sachs et al., 2022). This method was developed for individuals who identify as Hispanic, black, or white. In order to include all participants in the analysis, those who did not identify as Hispanic or black were assigned percentiles according to the normative data for those who identify as white (n=6 Asian, n=2 Native American, n=4 Unknown). Those with an education level of “Other”, “Some college”, and “Post college” were treated as “High School Education”, “Associate’s Degree”, and “Graduate Degree” respectively in the model. Normal cognition, MCI, and dementia were defined as >10^th^ percentile, ≤10^th^ percentile but >5^th^ percentile, and ≤5^th^ percentile respectively.

-Lifestyle factors: smoking history, alcohol intake in the prior year, and duration of walking with moderate intensity or greater per week as reported by the participant

-Physical examination was conducted by a trained research coordinator or nurse trainee and included examination of cranial nerves, gross motor power, Romberg sign, and sensation to pinprick, vibratory sense, and light touch in the fingers and toes.

-Depression symptoms were assessed with modifications of the Center for Epidemiologic Studies Depression Scale (mCES-D). SOMMA administers the 10-item version of CES-D and MYHAT assesses depression with a modification of the scale in which responses are binary (yes/no). Responses in SOMMA for each item were dichotomized whereby responses of “Much of the time” or “Most or all of the time” were considered a response of “yes” for a given item, or otherwise no. A depression symptom total score was then calculated for each participant.

-Blood pressure: seated systolic and diastolic blood pressure in mm of Hg were assessed on the date of the baseline visit for each parent study; values include mean over two trials (in SOMMA) or one-time assessment (in MYHAT).

-Prescribed medications were ascertained via review, by study staff, of each medication bottle if they were brought by participants to the study visit or were otherwise self-reported by the participant. A total count, number of any prescriptions reported, was determined. Intake of antidepressants (which could theoretically impact [^11^C]DTBZ binding)(Yasumoto et al., 2009) within 30 days of PET scan (SOMMA) or at the study visit closest to PET scan (MYHAT) was ascertained.

-Number of comorbidities: a count of 8 medical conditions, one point for each participant-reported condition: cancer, cardiac arrhythmia, chronic kidney disease, chronic obstructive pulmonary disease, coronary artery disease (myocardial infarction), congestive heart failure, diabetes, and stroke

The original study protocol aimed to assess pulmonary function with forced vital capacity but this was precluded during a portion of the COVID19 pandemic, hence this variable is not included.

#### 2.1.3 Statistical methods

The primary predictor was [^11^C]DTBZ BP_ND_ in each of the 3 striatal subregions (posterior putamen, anteroventral striatum, dorsal striatum). The primary outcome was %DGS. Gait speeds obtained during usual walking on even and uneven surface were examined as secondary outcomes.

PSRF that may affect gait speed included joint pain (in hip or leg), obesity, and/or reduced grip strength. The group with one or more PSRF that may affect gait speed is hereto forth referred to as the “any PSRF” group, and the group with absence of any PSRF as the “no PSRF” group.

Sample characteristics were summarized with descriptive statistics. The PSRF and no PSRF groups were compared with two-sample t-test, two-sample t-test with unequal variances, Wilcoxon rank sum test, chi-square test, or Fisher’s exact test as appropriate. Bivariate relationships between gait speed and age, and other demographic/clinical and imaging characteristics, all centered and standardized, were examined with simple linear regression models to obtain standardized beta coefficients. A similar approach was followed to examine the relationship between [^11^C]DTBZ BP_ND_ and age and [^11^C]DTBZ BP_ND_ and age.

To determine if there was a significant relationship between gait speed and [^11^C]DTBZ BP_ND_, simple linear regressions were conducted with %DGS as the outcome, [^11^C]DTBZ BP_ND_ in each of the 3 striatal subregions as predictors (each in separate models), and age at PET scan and sex as covariates. To examine whether the contribution of dopaminergic terminal integrity to %DGS varies according to presence or absence of any PSRF, an interaction term between DTBZ binding in each region and presence/absence of any PSRF was entered into the model, with age and sex as covariates. If the interaction was significant, models were repeated stratified by any PSRF and no PSRF.

Models where the main association was significant in the sample as a whole or in any stratum were adjusted for additional confounders. Given the high number of potential confounders, and to address potential collinearity, variables were entered in subsequent steps: (1) any variable determined to be possibly (p≤0.10) associated with the outcome in bivariate analysis; (2) log-transformed white matter hyperintensities; (3) taking antidepressant (4) diabetes and number of comorbidities. Adjusted R^2^ was used to determine percent of the variance in the outcome explained by the model.

Assumptions for linear regression were checked by examining normality of distribution of the residuals for each model. All analyses were conducted with Stata version 18 (StataCorp, College Station, Tx, USA). A p-value <0.05 was considered statistically significant and no adjustment was made for multiple comparisons.

## 3. Results

Among potential participants, 644 were screened and 237 gave consent to participate and completed some study assessments (Figure 1). Compared to those who consented, those who did not were older (73.94 (4.70) vs 75.39 years (5.94), p=0.0009) and were more likely to have a level of education no higher than a High school/General educational development (15.19% vs 24.65%, p=0.003). All those excluded due to missing gait measures were from the SOMMA cohort and were more likely to be male compared to those who had a gait assessment (71.43% vs 37.81%, p<0.001).

**Figure 1.**
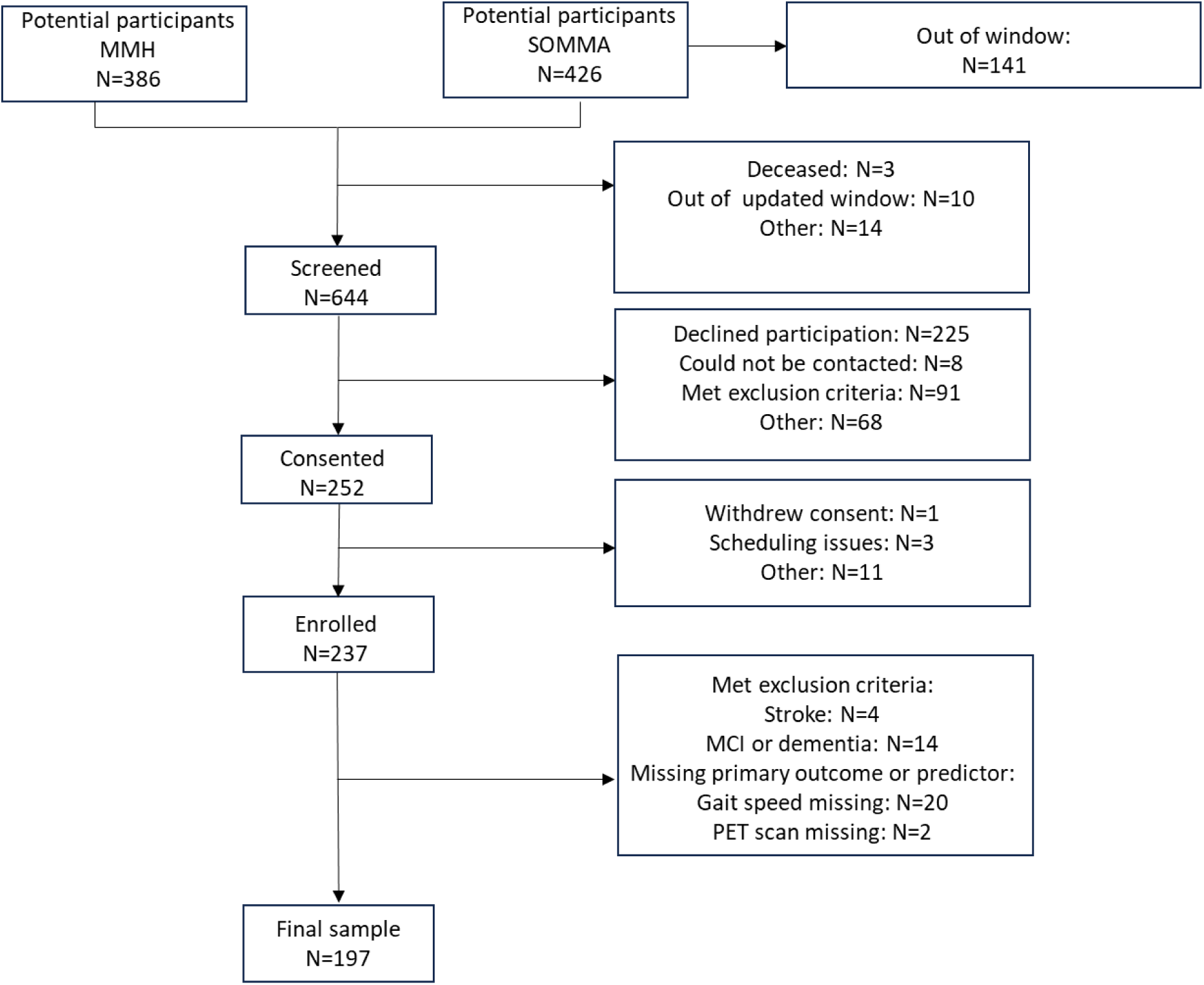
STROBE diagram for the study.

The final sample in the analysis included 197 participants. Characteristics of the final sample are shown in Table 1. Compared to the MYHAT cohort, the SOMMA cohort were older, had a higher level of education, were less likely to have smoked, were less likely to be obese, less likely to have diabetes, and had a greater burden of white matter hyperintensities(Supplementary Table 1). Differences between cohorts in %DGS or [^11^C]DTBZ BP_ND_ were not statistically significant.

**Table 1:** Cohort characteristics in the sample as a whole and in those without vs with any peripheral systemic risk factor (PSRF) for gait slowing.

| Variable<sup>i</sup> | Total sample<br>N=197 | No PSRF<br>N=67 | Any PSRF<br>N=130 | p-value<sup>ii</sup> |
| --- | --- | --- | --- | --- |
| Age (years) | 74.92 (4.53) | 75.06 (4.97) | 74.85 (4.30) | 0.76 |
| Sex |  |  |  | 0.02 |
| Male | 75 (38.07%) | 33 (49.25%) | 42 (32.31%) |  |
| Female | 122 (61.93%) | 34 (50.75%) | 88 (67.69%) |  |
| Self-Identified Race |  |  |  | 0.88 |
| White | 179 (90.86%) | 63 (94.03%) | 116 (89.23%) |  |
| Black | 15 (7.61%) | 4 (5.97%) | 11 (8.46%) |  |
| Asian | 1 (0.51%) | 0 (0%) | 1 (0.77%) |  |
| Native American/Alaskan Native | 1 (0.51%) | 0 (0%) | 1 (0.77%) |  |
| Unknown | 1 (0.51%) | 0 (0%) | 1 (0.77%) |  |
| Education |  |  |  | 0.005 |
| Highschool/GED | 32 (16.24%) | 4 (5.97%) | 28 (21.54%) |  |
| More than Highschool | 164 (83.25%) | 63 (94.03%) | 101 (77.69%) |  |
| Other | 1 (0.51%) | 0 (0%) | 1 (0.77%) |  |
| Smoking status |  |  |  | 0.25 |
| Never | 105 (53.30%) | 41 (61.19%) | 64 (49.23%) |  |
| Past | 80 (40.61%) | 22 (32.84%) | 58 (44.62%) |  |
| Current | 12 (6.09%) | 4 (5.97%) | 8 (6.15%) |  |
| Body mass index (kg/m<sup>2</sup>) | 28.52 (5.45) | 25.67 (2.67) | 29.98 (5.93) | <0.001 |
| Alcohol Intake Past Year |  |  |  | 0.60 |
| No | 48 (24.62%) | 18 (26.87%) | 30 (23.44%) |  |
| Yes | 147 (75.38%) | 49 (73.13%) | 98 (76.56%) |  |
| Physical Activity: Hours Walked/Week | 1.00 (0.00-3.50) | 1.50 (0.00-3.50) | 0.58 (0.00-3.50) | 0.45 |
| Depression Symptoms | 0.00 (0.00-1.00) | 0.00 (0.00-1.00) | 0.00 (0.00-1.00) | 0.34 |
| Diabetes<sup>iii</sup> |  |  |  | 0.44 |
| No | 161 (82.14%) | 57 (85.07%) | 104 (80.62%) |  |
| Yes | 35 (17.86%) | 10 (14.93%) | 25 (19.38%) |  |
| Systolic Blood Pressure<sup>iii</sup> | 131.41 (16.07) | 130.98 (16.95) | 131.65 (15.64) | 0.80 |
| Diastolic Blood Pressure<sup>iii</sup> | 71.78 (8.82) | 71.97 (8.78) | 71.68 (8.88) | 0.84 |
| Pulse | 65.74 (9.80) | 64.42 (8.88) | 66.46 (10.24) | 0.20 |
| Medication Count<sup>ii</sup> | 4.00 (2.00-6.00) | 3.00 (1.00-5.00) | 5.00 (3.00-7.00) | <0.001 |
| Treated with antidepressant | 37 (18.78) | 8 (11.94) | 29 (22.31) | 0.08 |
| Number of comorbidities |  |  |  | 0.29 |
| 0 | 97 (49.49%) | 36 (53.73%) | 61 (47.29%) |  |
| 1 | 78 (39.80%) | 27 (40.30%) | 51 (39.53%) |  |
| 2 or 3 | 21 (10.71%) | 4 (5.97%) | 17 (13.18%) |  |
| Positive Romberg sign<sup>ii</sup> |  |  |  | -- |
| No | 194 (100%) | 66 (100%) | 128 (100%) |  |
| Yes | 0 | 0 | 0 |  |
| Vibratory sense reduced in toes<sup>iii</sup> |  |  |  | 1.00 |
| No | 77 (39.69%) | 27 (40.30%) | 50 (39.37%) |  |
| Yes | 117 (60.31%) | 40 (59.70%) | 77 (60.63%) |  |
| Pinprick sensation reduced in toes<sup>iii</sup> |  |  |  | 0.66 |
| No | 189 (97.42%) | 66 (98.51%) | 123 (96.85%) |  |
| Yes | 5 (2.58%) | 1 (1.49%) | 4 (3.15%) |  |
| Light touch sensation reduced<sup>iii</sup> |  |  |  | 1.00 |
| No | 192 (98.46%) | 66 (98.51%) | 126 (98.44%) |  |
| Yes | 3 (1.54%) | 1 (1.49%) | 2 (1.56%) |  |
| White matter hyperintensities (Normalized, mm<sup>3</sup> <sup>iii</sup>) | 0.007 (0.004-0.015) | 0.007 (0.004-0.017) | 0.007 (0.004-0.015) | 0.99 |
| Gray matter atrophy | 0.336 (0.027) | 0.338 (0.029) | 0.335 (0.026) | 0.39 |
| Subcortical gray matter atrophy | 0.034 (0.003) | 0.034 (0.003) | 0.034 (0.004) | 0.36 |
| Low Grip Strength<sup>iii</sup> |  |  |  | -- |
| No | 135 (69.95%) | 65 (100%) | 70 (54.69%) |  |
| Yes | 58 (30.05%) | 0 (0%) | 58 (45.31%) |  |
| Obese (body mass index≥30 kg/m<sup>2</sup>) |  |  |  | -- |
| No | 128 (64.97%) | 67 (100%) | 61 (46.92%) |  |
| Yes | 69 (35.03%) | 0 (0%) | 69 (53.08%) |  |
| Pain for ≥1 month in hip or leg<sup>iii</sup> |  |  |  | -- |
| No | 137 (69.54%) | 67 (100%) | 70 (53.85%) |  |
| Yes | 60 (30.46%) | 0 (0%) | 60 (46.15%) |  |
| Number of peripheral systemic risk factors for gait slowing<sup>iv</sup> |  |  |  | -- |
| 0 | 67 (34.01%) | 67 (100%) | 0 (0%) |  |
| 1 | 77 (39.09%) | 0 (0%) | 77 (59.23%) |  |
| 2 | 49 (24.87%) | 0 (0%) | 49 (37.69%) |  |
| 3 | 4 (2.03%) | 0 (0%) | 4 (3.08%) |  |
| Gait speed measures (m/s) |  |  |  |  |
| Usual Walking Speed Even Surface | 1.10 (0.20) | 1.17 (0.20) | 1.06 (0.19) | <0.001 |
| Usual Walking Speed Uneven Surface | 1.03 (0.21) | 1.12 (0.20) | 0.99 (0.20) | <0.001 |
| % Difference in gait speed uneven vs even (%DGS) | -5.37 (-9.34 to -3.00) | -5.03 (-6.73 to -1.95) | -5.77 (-10.06 to -3.27) | 0.05 |
| [<sup>11</sup>C]DTBZ BP<sub>ND</sub> |  |  |  |  |
| Posterior putamen (sensorimotor striatum) | 2.44 (0.42) | 2.38 (0.39) | 2.46 (0.43) | 0.20 |
| Anteroventral striatum (limbic striatum) | 1.63 (0.27) | 1.62 (0.24) | 1.64 (0.28) | 0.60 |
| Caudate and anterior putamen (associative striatum) | 2.08 (0.34) | 2.04 (0.29) | 2.11 (0.35) | 0.16 |

<sup>i</sup>values shown are N (%), mean (SD), or median (interquartile range)
<sup>ii</sup> p-value from results comparing any PSRF to no PSRF groups with two-sample t-test, two-sample t-test with unequal variance, Wilcoxon rank sum test, chi-square test, or Fisher's exact test as appropriate.
<sup>iii</sup> Missing values: Romberg n=3, vibratory sense n=3, light touch sensation n=3, alcohol intake n=2, physical activity n=6, diabetes assessment n=1, BP measurement n=27, WHITE MATTER HYPERINTENSITIES n=4, grip strength n=4, prescription count n=1
<sup>iv</sup>All 3 systemic risk factors assessed in n=193; 2 impairments assessed in n=4. In 2 participants no impairments were present but not all 3 risk factors were assessed
DTBZ BP<sub>ND</sub> =dihydrotetrabenazine binding potential, GED=general education diploma . PSRF=peripheral systemic risk factors.

In the sample as a whole, age was inversely associated with the [^11^C]DTBZ BP_ND_ in the posterior putamen, anteroventral striatum, dorsal striatum (standardized β = -0.234, p=0.001, standardized β = -0.242, p=0.001, -standardized β = -0.197, p=0.006 respectively). Age was not associated with %GCS (standardized β =0.094, p=0.188). On average, %DGS was median -5.37 (interquartile range (IQR) -9.34 to -3.0).

Peripheral systemic risk factors for gait slowing were present in 130 (65.99%) and no PSRF were present in 67 (34.01%) (Table 1). Compared to the no PSRF, the any PSRF were more likely to be female, to have lower education, higher mean BMI (as expected given absence of obesity in the no PSRF group by definition), and had a greater number of prescribed medications. Other characteristics did not significantly differ between groups. The group with any PSRF slowed down more on transition to uneven surface compared to the group with no PSRF (median (IQR) -5.77 (- 10.06- to -3.27) vs -5.03 (-6.73 to -1.95) respectively), p=0.05). Other factors possibly (p≤0.10) associated with %DGS in the sample as a whole included sex, race, education, physical activity, and medication count (Table 2).

**Table 2.** Relationship between difference in gait speed from even to uneven surface and covariates/possible confounders in the cohort as a whole and in those with any peripheral systemic risk factors (PSRF) compared to those with no PSRF

| Variables (standardized) | Total sample (n=197) |  |  | Any PSRF (n=130) |  |  | No PSRF (n=67) |  |  |
| --- | --- | --- | --- | --- | --- | --- | --- | --- | --- |
| | s$\beta^i$ | SE | p-value | s$\beta^i$ | SE | p-value | s$\beta^i$ | SE | p-value |
| Age | 0.09 | 0.07 | 0.19 | 0.17 | 0.10 | 0.09 | -0.03 | 0.08 | 0.73 |
| Female sex | -0.38 | 0.15 | 0.01 | -0.45 | 0.20 | 0.03 | -0.14 | 0.18 | 0.42 |
| Identified as non-White Race or Other | -1.01 | 0.24 | <0.001 | -1.33 | 0.29 | <0.001 | 0.23 | 0.37 | 0.54 |
| Education, Highschool/GED reference |  |  |  |  |  |  |  |  |  |
| More than Highschool/Other | 0.39 | 0.19 | 0.05 | 0.25 | 0.24 | 0.30 | 0.71 | 0.36 | 0.05 |
| Smoking, Never reference |  |  |  |  |  |  |  |  |  |
| Past | 0.18 | 0.15 | 0.23 | 0.19 | 0.20 | 0.35 | 0.29 | 0.19 | 0.13 |
| Current | -0.05 | 0.31 | 0.88 | -0.08 | 0.42 | 0.84 | 0.08 | 0.37 | 0.82 |
| Alcohol Intake Past Year | 0.06 | 0.16 | 0.73 | 0.07 | 0.23 | 0.74 | 0.06 | 0.20 | 0.76 |
| Physical Activity: Hours Walked/Week | 0.16 | 0.07 | 0.03 | 0.16 | 0.09 | 0.09 | 0.15 | 0.10 | 0.14 |
| Depression Symptoms | -0.01 | 0.07 | 0.92 | 0.02 | 0.09 | 0.86 | 0.00 | 0.15 | 0.99 |
| Diabetes | -0.29 | 0.19 | 0.12 | -0.34 | 0.25 | 0.17 | -0.11 | 0.25 | 0.67 |
| Systolic BP | 0.03 | 0.08 | 0.72 | 0.12 | 0.11 | 0.30 | -0.11 | 0.08 | 0.17 |
| Diastolic BP | -0.13 | 0.08 | 0.11 | -0.16 | 0.11 | 0.14 | -0.06 | 0.08 | 0.43 |
| Medication Count | -0.19 | 0.07 | 0.01 | -0.22 | 0.10 | 0.03 | 0.01 | 0.10 | 0.95 |
| Treated with antidepressant | -0.19 | 0.18 | 0.30 | -0.18 | 0.23 | 0.45 | -0.03 | 0.27 | 0.91 |
| Number of comorbidities, 0 reference |  |  |  |  |  |  |  |  |  |
| 1 | 0.03 | 0.15 | 0.83 | 0.10 | 0.21 | 0.62 | -0.07 | 0.19 | 0.69 |
| 2 or 3 | -0.23 | 0.24 | 0.34 | -0.21 | 0.30 | 0.49 | 0.02 | 0.38 | 0.95 |
| Vibratory sense reduced in toes | 0.01 | 0.15 | 0.97 | 0.10 | 0.20 | 0.61 | -0.17 | 0.18 | 0.35 |
| Pinprick sensation reduced in toes | -0.43 | 0.46 | 0.34 | -0.25 | 0.57 | 0.65 | -0.89 | 0.72 | 0.22 |
| Light touch sensation reduced | -0.13 | 0.58 | 0.83 | -0.39 | 0.79 | 0.62 | 0.41 | 0.72 | 0.57 |
| Log-transformed white matter hyperintensities | -0.06 | 0.06 | 0.31 | -0.12 | 0.09 | 0.21 | 0.02 | 0.07 | 0.82 |
| Parent Study (SOMMA or MYHAT) | 0.09 | 0.15 | 0.52 | 0.09 | 0.20 | 0.64 | 0.02 | 0.18 | 0.92 |
| Gray matter atrophy | -0.08 | 0.07 | 0.28 | -0.09 | 0.10 | 0.38 | -0.08 | 0.08 | 0.33 |
| Subcortical gray matter atrophy | -0.08 | 0.07 | 0.29 | 0.10 | 0.20 | 0.61 | -0.09 | 0.09 | 0.35 |

Each row represents results of a separate linear regression model
GED=general education diploma . PSRF=peripheral systemic risk factors. $s\beta$ =standardized beta coefficient. WHITE MATTER
HYPERINTENSITIES=white matter hyperintensity

In the sample as a whole, [^11^C]DTBZ BP_ND_ in none of the 3 striatal subregions were associated with %DGS after adjusting for age and sex (Table 3). When an interaction term between presence/absence of any PSRF for gait slowing and [^11^C]DTBZ BP_ND_ in each region was introduced into the model, the interaction was significant for posterior putamen only (p=0.03) but not for other subregions (Table 3; Figure 2)), indicating that the effect of posterior putamen on gait speed is influenced by the presence or absence of PSRF.

**Figure 2.**
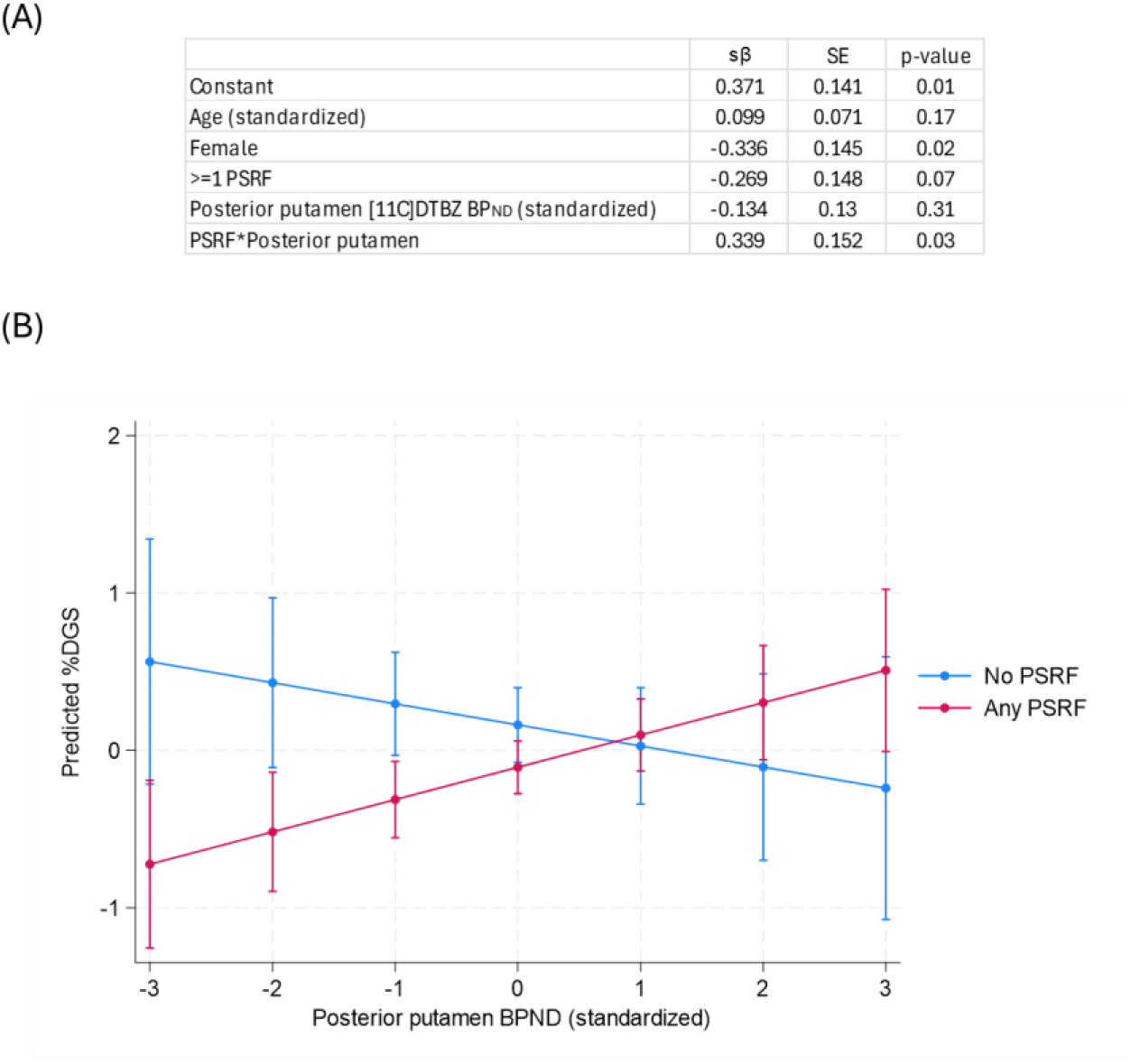
(A) Full results of regression model examining interaction between PSRF and posterior putamen on % difference in gaits speed, adjusting for age and sex and (B) graphical depiction of interaction between presence of PSRF and posterior putamen when holding age and sex constant

**Table 3.** Relationship between difference in gait speed on transition from even to uneven surface and DTBZ binding in the sample as a whole and interaction with presence or absence of any peripheral systemic risk factor for gait impairment, adjusted for age and sex

|  | Total Sample<sup>i</sup> |  |  | Interaction term |
| --- | --- | --- | --- | --- |
|  | sβ | SE | p-value | p-value<sup>ii</sup> |
| [<sup>11</sup>C]DTBZ BP<sub>ND</sub> in striatal region |  |  |  |  |
| Posterior putamen (sensorimotor striatum) | 0.01 | 0.07 | 0.18 | 0.03 |
| Anteroventral striatum (limbic striatum) | 0.02 | 0.08 | 0.77 | 0.29 |
| Caudate and anterior putamen (associative striatum) | 0.10 | 0.07 | 0.19 | 0.06 |

<sup>i</sup> Linear regression model consisting of: difference in gait speed on transition from even to uneven surface (outcome), [<sup>11</sup>C]DTBZ BP<sub>ND</sub> in specified striatal region, age, and sex
<sup>ii</sup> Linear regression model consisting of: difference in gait speed on transition from even to uneven surface (outcome), [<sup>11</sup>C]DTBZ BP<sub>ND</sub> in specified striatal region, age, and sex, presence of any risk factor, and interaction term between risk factor present (yes/no) and [<sup>11</sup>C]DTBZ BP<sub>ND</sub> in specified striatal region
DTBZ BP<sub>ND</sub> =dihydrotetrabenazine binding potential. PSRF=peripheral systemic risk factors. sβ=standardized beta coefficient. SE=standard error

Accordingly, we proceeded to further examine the relationship between %DGS and posterior putamen [^11^C]DTBZ BP_ND_ in the any PSRF group. Associations between usual gait speeds on even or uneven surface and [^11^C]DTBZ BP_ND_ in each of the striatal subregions were not significant and the interaction terms were not significant (Supplementary Table 2).

Among the group with any PSRF for gait slowing, posterior putamen [^11^C]DTBZ BP_ND_ was significantly associated with %DGS, independent of age and sex (Table 4, Model 1). The association was also independent of confounders selected using bivariate models (Table 3): race, amount of moderate walking per week, medication count, and education (Table 4, Model 2). In addition, the association remained independent when intake of antidepressant (which could affect [^11^C]DTBZ BP_ND_) was added to the model (Model 3). However, when other confounders selected a priori (white matter hyperintensities, number of comorbidities,) were adjusted for, the relationship between posterior putamen and %DGS was no longer significant (Table 4, models 4-6). Model 2 explained about 21.8% of the variance of %DGS. In this model, when adjusting for covariates, the variables that were significantly (p<0.05) and independently associated with higher %DGS (less slowing down) in those with any PSRF were higher posterior putamen DTBZ BP_ND,_ younger age, identifying as white race, and lower number of prescribed medications.

**Table 4:**
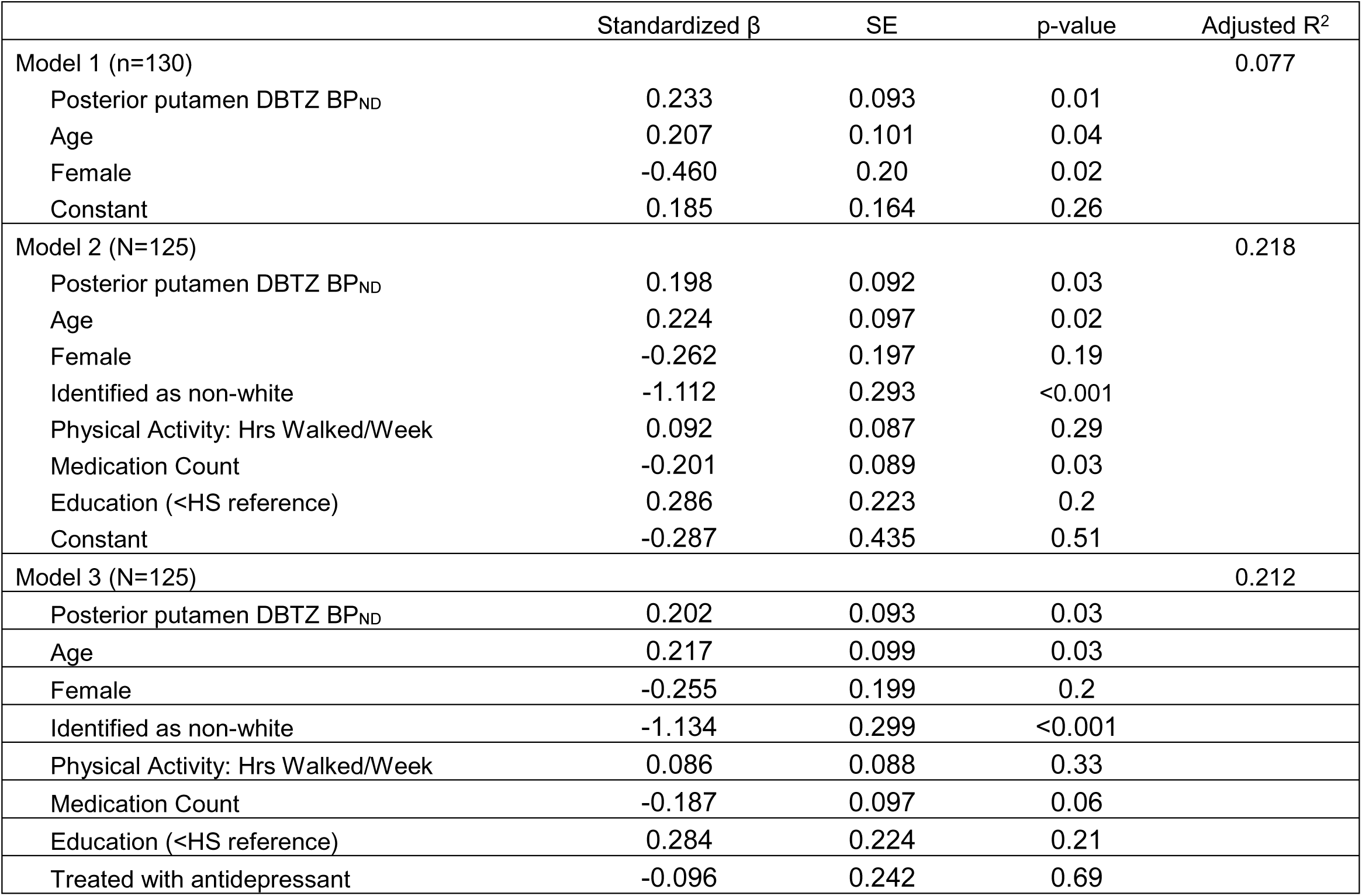
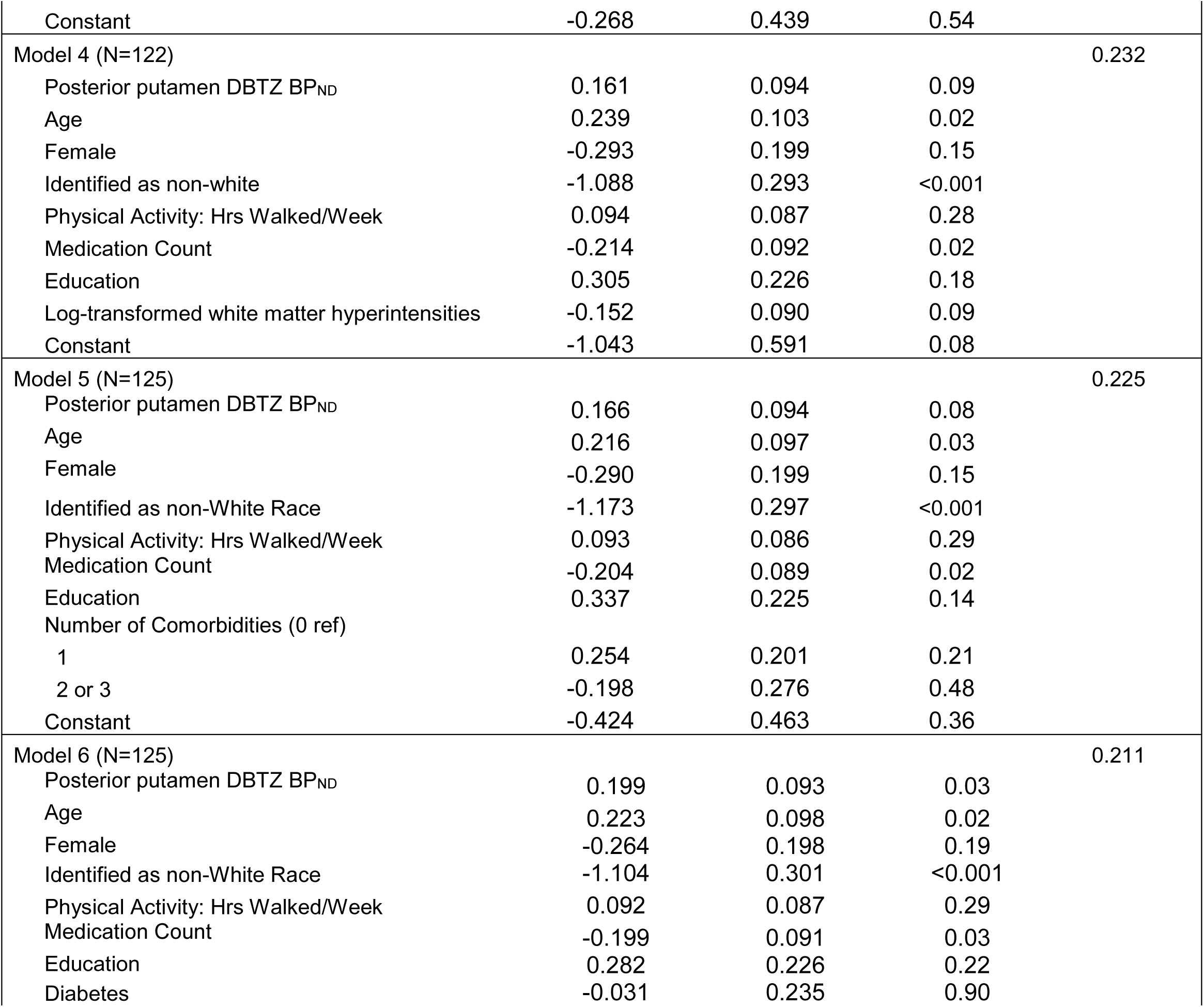
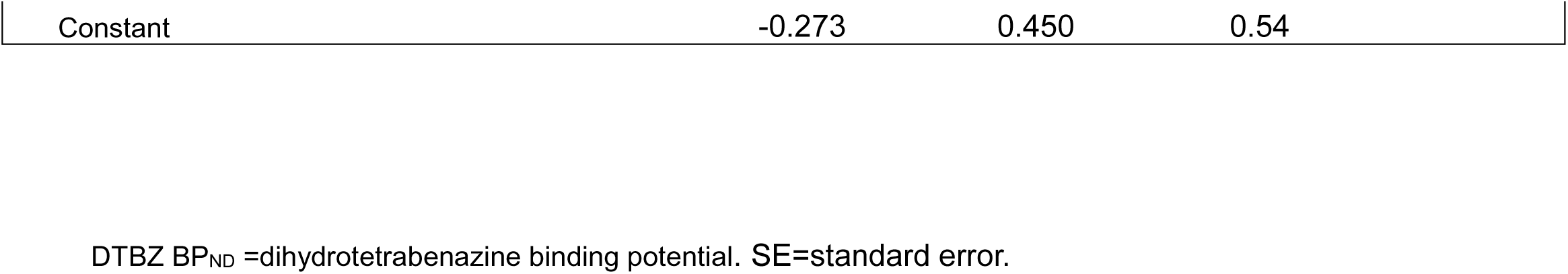
Among those with any peripheral systemic risk factor for gait slowing (n=130), results of multivariable regression with % difference in gait speed on transition from uneven to even surface as outcome and PET-derived index of VMAT2 availability in the posterior putamen as the predictor.

| | Standardized $\beta$ | SE | p-value | Adjusted R<sup>2</sup> |
| --- | --- | --- | --- | --- |
| Model 1 (n=130) |  |  |  | 0.077 |
| Posterior putamen DBTZ BP<sub>ND</sub> | 0.233 | 0.093 | 0.01 |  |
| Age | 0.207 | 0.101 | 0.04 |  |
| Female | -0.460 | 0.20 | 0.02 |  |
| Constant | 0.185 | 0.164 | 0.26 |  |
| Model 2 (N=125) |  |  |  | 0.218 |
| Posterior putamen DBTZ BP<sub>ND</sub> | 0.198 | 0.092 | 0.03 |  |
| Age | 0.224 | 0.097 | 0.02 |  |
| Female | -0.262 | 0.197 | 0.19 |  |
| Identified as non-white | -1.112 | 0.293 | <0.001 |  |
| Physical Activity: Hrs Walked/Week | 0.092 | 0.087 | 0.29 |  |
| Medication Count | -0.201 | 0.089 | 0.03 |  |
| Education (<HS reference) | 0.286 | 0.223 | 0.2 |  |
| Constant | -0.287 | 0.435 | 0.51 |  |
| Model 3 (N=125) |  |  |  | 0.212 |
| Posterior putamen DBTZ BP<sub>ND</sub> | 0.202 | 0.093 | 0.03 |  |
| Age | 0.217 | 0.099 | 0.03 |  |
| Female | -0.255 | 0.199 | 0.2 |  |
| Identified as non-white | -1.134 | 0.299 | <0.001 |  |
| Physical Activity: Hrs Walked/Week | 0.086 | 0.088 | 0.33 |  |
| Medication Count | -0.187 | 0.097 | 0.06 |  |
| Education (<HS reference) | 0.284 | 0.224 | 0.21 |  |
| Treated with antidepressant | -0.096 | 0.242 | 0.69 |  |
| Constant | -0.268 | 0.439 | 0.54 |  |
| Model 4 (N=122) |  |  |  | 0.232 |
| Posterior putamen DBTZ BP<sub>ND</sub> | 0.161 | 0.094 | 0.09 |  |
| Age | 0.239 | 0.103 | 0.02 |  |
| Female | -0.293 | 0.199 | 0.15 |  |
| Identified as non-white | -1.088 | 0.293 | <0.001 |  |
| Physical Activity: Hrs Walked/Week | 0.094 | 0.087 | 0.28 |  |
| Medication Count | -0.214 | 0.092 | 0.02 |  |
| Education | 0.305 | 0.226 | 0.18 |  |
| Log-transformed white matter hyperintensities | -0.152 | 0.090 | 0.09 |  |
| Constant | -1.043 | 0.591 | 0.08 |  |
| Model 5 (N=125) |  |  |  | 0.225 |
| Posterior putamen DBTZ BP<sub>ND</sub> | 0.166 | 0.094 | 0.08 |  |
| Age | 0.216 | 0.097 | 0.03 |  |
| Female | -0.290 | 0.199 | 0.15 |  |
| Identified as non-White Race | -1.173 | 0.297 | <0.001 |  |
| Physical Activity: Hrs Walked/Week | 0.093 | 0.086 | 0.29 |  |
| Medication Count | -0.204 | 0.089 | 0.02 |  |
| Education | 0.337 | 0.225 | 0.14 |  |
| Number of Comorbidities (0 ref) |  |  |  |  |
| 1 | 0.254 | 0.201 | 0.21 |  |
| 2 or 3 | -0.198 | 0.276 | 0.48 |  |
| Constant | -0.424 | 0.463 | 0.36 |  |
| Model 6 (N=125) |  |  |  | 0.211 |
| Posterior putamen DBTZ BP<sub>ND</sub> | 0.199 | 0.093 | 0.03 |  |
| Age | 0.223 | 0.098 | 0.02 |  |
| Female | -0.264 | 0.198 | 0.19 |  |
| Identified as non-White Race | -1.104 | 0.301 | <0.001 |  |
| Physical Activity: Hrs Walked/Week | 0.092 | 0.087 | 0.29 |  |
| Medication Count | -0.199 | 0.091 | 0.03 |  |
| Education | 0.282 | 0.226 | 0.22 |  |
| Diabetes | -0.031 | 0.235 | 0.90 |  |

|  |  |  |  |
| --- | --- | --- | --- |
| Constant | -0.273 | 0.450 | 0.54 |

DTBZ BP<sub>ND</sub> =dihydratetrabenazine binding potential. SE=standard error.

## 4. Discussion

We found that cross-sectionally, the relationship between striatal dopaminergic terminal integrity and slowing down of gait on transition from an even to an uneven surface varied according to presence or absence of a PSRF. Importantly, this relationship was found for %DGS but not walking speeds on an even or uneven surface. Among the 3 striatal subregions examined, this relationship was only present for the posterior putamen, consistent with the increased demand on sensorimotor integration required for complex walking tasks(Tian et al., 2017). Change in walking speed on surface transition is a possible measure of real-world perturbations in walking speed as may be encountered by an older adult navigating ground where the surface could change unpredictably from even to uneven(Menant et al., 2009; Menz et al., 2003; Zukowski et al., 2024), such as on a sidewalk. Slowing down on surface transitions could be more predictive of fall risk(Wenzel et al., 2023) and underlying neuropathology(Zukowski et al., 2024) compared to other gait measures. While posterior putamen [^11^C]DTBZ BP_ND_ was not associated with gait speed measures in the sample as a whole, in the group with a PSRF, there was a significant relationship with %DGS.

Only among individuals with joint pain, obesity, and/or low grip strength, greater nigral dopaminergic terminal integrity was associated with less slowing down during the transition from even to uneven surface. To date, several lines of evidence have indicated a relationship between the dopaminergic system and gait abnormalities in older adults without diagnosed neurologic diseases. The catechol-o-methyl transferase genotype, an enzyme that metabolizes dopamine and that is highly expressed in the prefrontal cortex, has consistently been associated with gait speed cross-sectionally predicts decline in gait speed in older adults(Mance et al., 2020; Metti et al., 2017; Sprague et al., 2021). Recognizing that catechol-o-methyl transferase genotype is only an indirect reflection of CNS dopaminergic neurotransmission, and that it predominantly may influence prefrontal cortex and not striatal dopamine and other catecholamines, additional data were garnered from imaging studies in healthy adults without diagnosed neurologic diseases. Reductions in binding of PET radioligands for the dopamine transporter (DAT) have been associated with reduced gait speed(Cham et al., 2008; Rosano et al., 2019). Similar investigations have provided evidence for the potential of dopaminergic neurotransmission to provide resilience against central and peripheral contributors to gait impairment. In one study(Rosso et al., 2018), while catechol-o-methyl transferase genotype was not associated with walking speed, the genotype modified the relationship between white matter hyperintensities and gait speed. Specifically, an association between white matter hyperintensities and gait speed was present in homozygous or heterozygous Val carriers but not Met/Met homozygotes(Rosso et al., 2018). These results were interpreted as higher levels of prefrontal cortex dopamine in those with Met/Met genotype conferring compensation for white matter hyperintensities on gait speed(Rosso et al., 2018). In addition, striatal DAT binding was shown to attenuate the relationship between white matter hyperintensities and parkinsonian signs(Rosano et al., 2019).

Nigrostriatal dopaminergic neurotransmission regulates the automated execution of overlearned motor tasks via its connections with sensorimotor cortical and subcortical areas(Wu and Hallett, 2005). Peripheral systemic risk factors for gait slowing such as joint pain, obesity, or reduced muscle strength make walking more demanding, eventually exceeding the capacity of the sensorimotor network for habitual control of overlearned movements. In such individuals with PSRF, the networks that modulate gait may call on the dopaminergic system to help compensate for the PSRF via higher sensorimotor connectivity, thus increasing automaticity of walking. That is, a stronger sensorimotor control system could offer at least some resilience to the demands imposed by PSRF, and promoting walking performance in a taxing environment such as transitions from even to uneven surfaces. Consistent with this, we found a relationship between posterior putamen, the sensorimotor region of the striatum, but not the anteroventral striatum which mainly projects to the limbic regions or associative striatum which mainly projects to the prefrontal cortex. In a mouse model, higher nigrostriatal dopaminergic levels had a net effect of facilitating striato-pallidum-thalamic excitatory outputs to primary sensorimotor regions, promoting ease and fluidity of movements, and initiation and execution of coordinated sequences of musculoskeletal activations(Panigrahi et al., 2015). Preliminary data supporting this model in humans comes from studies in Parkinson’s disease, where dopaminergic neurotransmission was associated with microstructural and functional connectivity of sensorimotor networks(Carbonell et al., 2014; Gilat et al., 2017; Manza et al., 2016; Nagano-Saito et al., 2017; Szewczyk-Krolikowski et al., 2014; Yang et al., 2016). In future studies we will pursue mechanistic investigations utilizing functional connectivity imaging to further test this model in older adults without diagnosed neurologic disorders.

Posterior putamen [^11^C]DTBZ BP_ND_ and %DGS were independently associated among those with PSRF, and this relationship remained significant after adjustment for several possible confounders. Older age was inversely associated with [^11^C]DTBZ BP_ND_, as expected given the established age-dependent decline in dopaminergic neurotransmission(Cham et al., 2008). However, in those with PSRF, the association of [^11^C]DTBZ BP_ND_ with %DGS was independent of age, indicating the influence of dopaminergic neurotransmission of gait speed is not entirely explained by older age. When adjusting (in separate models) for race, physical activity, medications, and education the relationship remained similar, as it did for diabetes. However, when adjusting for cerebral small vessel disease or number of comorbidities, the relationship was no longer significant. This may suggest that with these additional CNS or systemic risk factors, dopaminergic neurotransmission is no longer able to offer resilience to effects on slowing of gait on transition. On the other hand, the findings may reflect collinearity between medical comorbidities and white matter hyperintensities measures, or medication count and comorbidities respectively.

Due to the protocol limitations imposed during the COVID19 pandemic in 2020-2021, we were unable to assess pulmonary function, a known contributor to slow gait speed. Other contributors to slow gait speed that were not assessed in our study include (but are not limited to) vision impairment, peripheral vascular disease, atherosclerotic vascular disease, and blood-based measures of glucose dysmetabolism and renal dysfunction(Rosso et al., 2015). Thus, it is not surprising that our model only explained at most about one-fifth of the variance in change in gait speed on transition from even to uneven surface. These findings reflect the multifactorial nature of gait speed slowing in older adults.

Strengths of this study include a comprehensive assessment of risk factors for gait slowing including a neuroimaging protocol to assess for cerebral small vessel disease and GMV acquired simultaneously with PET. There are several limitations to this study. This was a cross-sectional study and does not allow us to determine causality—that is, to prove that it was dopaminergic neuron integrity that confers resilience to PSRF in older adults. In addition, only 30% of screened participants were included in the final analytic sample, and our study sample lacks racial and ethnic diversity. This is not uncommon in neuroimaging studies of older adults. Thus, our results may not be generalizable to the older adult population from which our sample was drawn. In addition, as mentioned, we only assessed 3 PSRF that may impact gait. Furthermore, while we were able to account for the possible effect of antidepressant intake on [^11^C]DTBZ BP_ND_, there are other medications such as stimulants which could impact VMAT2 binding that we did not have data for(Eiden and Weihe, 2011; Narendran et al., 2012; Yasumoto et al., 2009).

In conclusion, in this cross-sectional study of older adults, among those with joint pain, reduced grip strength, and/or obesity, greater dopaminergic neuron terminal integrity was significantly associated with less gait slowing when transitioning from an even to uneven surface. These results may inform future interventions to ameliorate gait performance by increasing dopamine in older adults with PSRF for gait impairment.

## Data Availability

All data is publicly available

https://www.sommastudy.com/

## Abbreviations

MYHAT: Monongahela-Youghiogheny Healthy Aging Team study
PET: Positron Emission Tomography
PSRF: Peripheral Systemic Risk Factors
SOMMA: Study of Muscle, Mobility and Aging
VMAT2: type-2 vesicular monoamine transporter

## Author roles

Chahine LM: Formal analysis, Writing - original draft

Rosso A: Methodology, Writing - review & editing.

Troidl I: Project administration, Data curation, Data Validation, Writing - review & editing.

Ganguli M: Writing - Methodology, review & editing.

Newman: Writing - Funding acquisition, Methodology, review & editing.

Cummings S: Funding acquisition Writing - review & editing.

Studenski S: Writing - review & editing.

Sparto PJ: Writing - review & editing.

Lopresti B: Methodology, Writing - review & editing.

Royse S: Methodology, Writing - review & editing.

Huppert T: Methodology, Writing - review & editing.

Redfern M: Writing - review & editing.

Sparto PJ: Writing - review & editing.

Bohnen N: Methodology, Funding acquisition, Writing - review & editing.

Rosano C: Formal analysis, Funding acquisition, Methodology, Supervision, Writing - original draft

## Declaration of Generative AI and AI-assisted technologies

No generative AI or AI-assisted technologies were used in the writing process.

---

**Supplementary Table 1.**
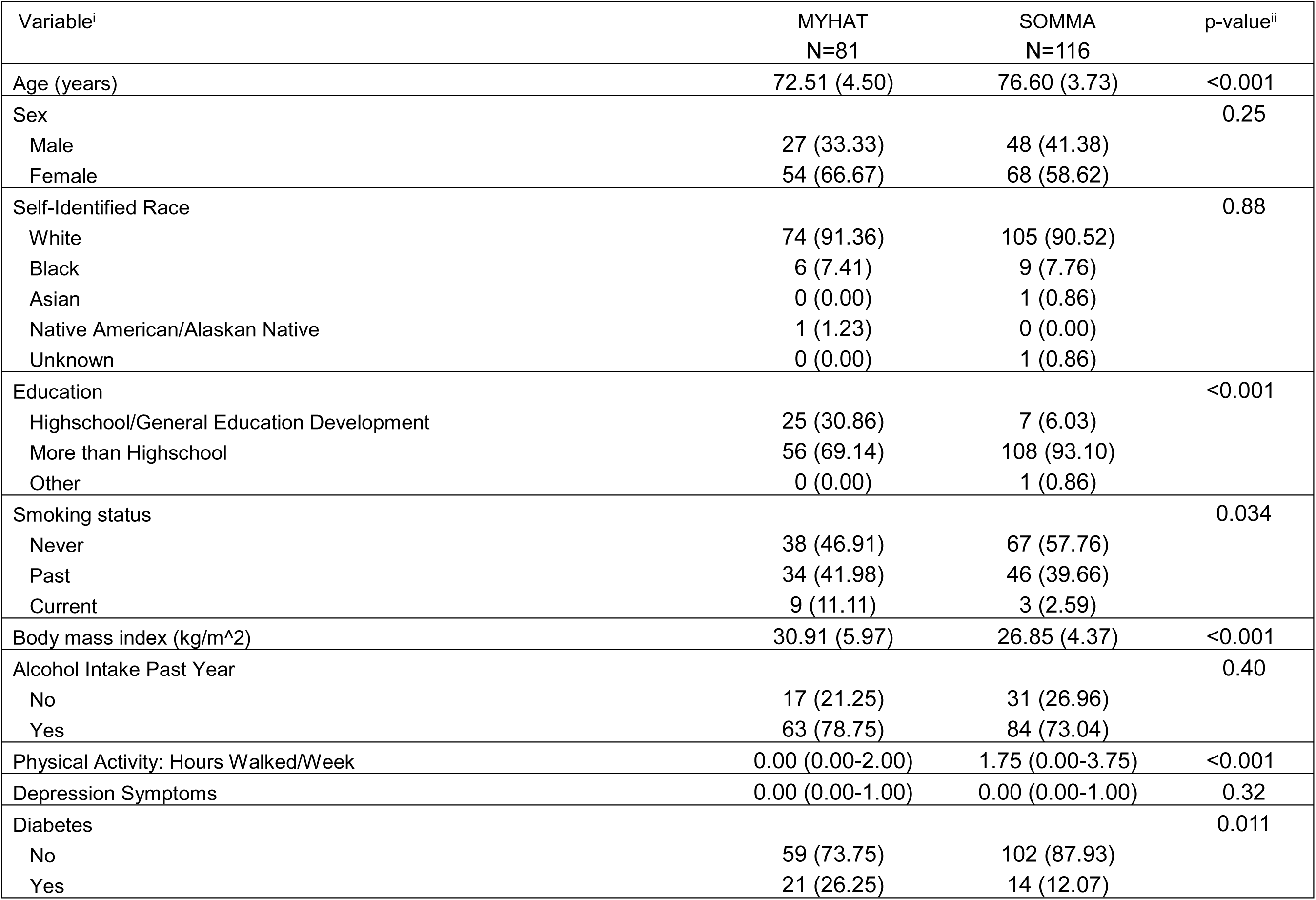
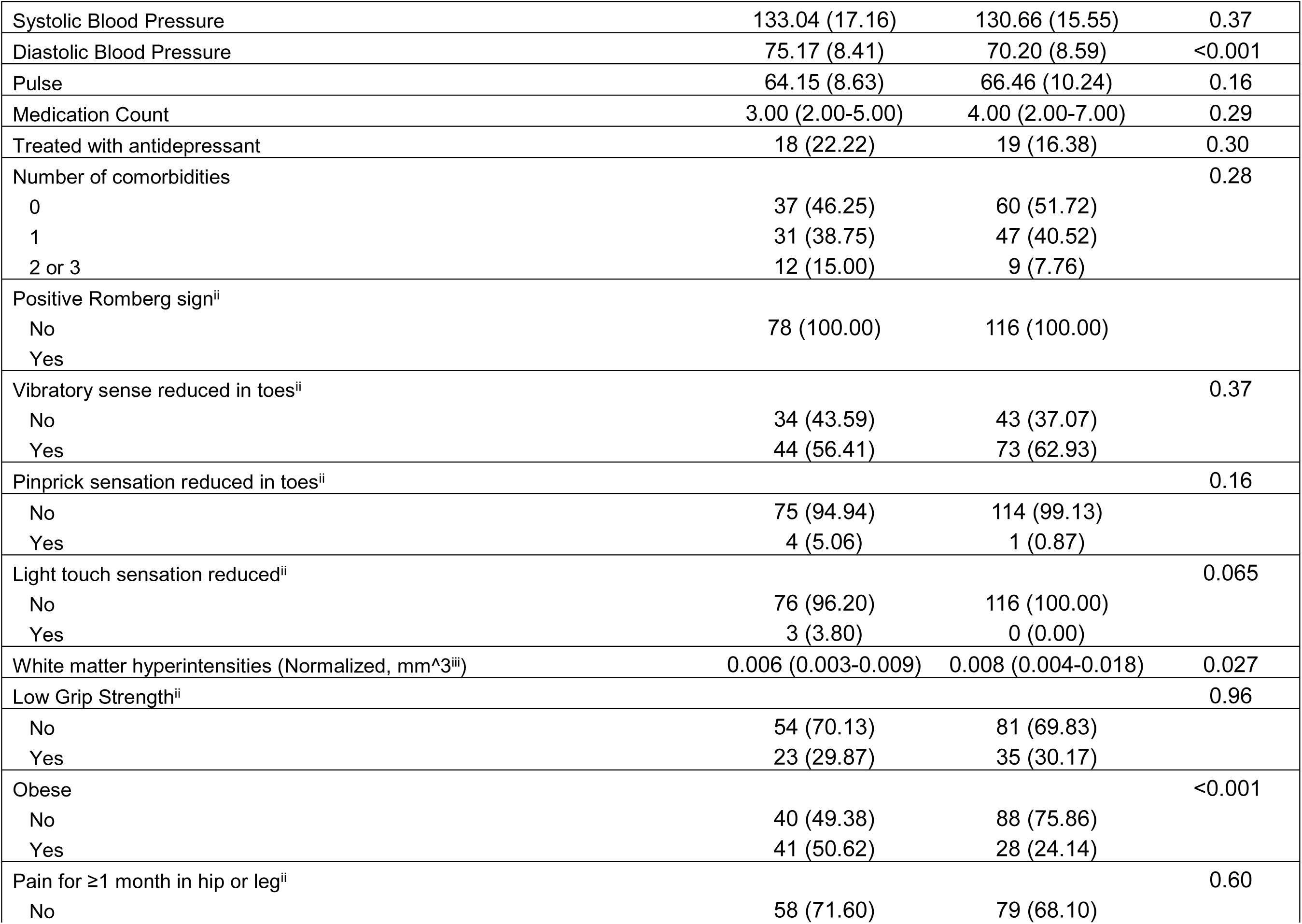
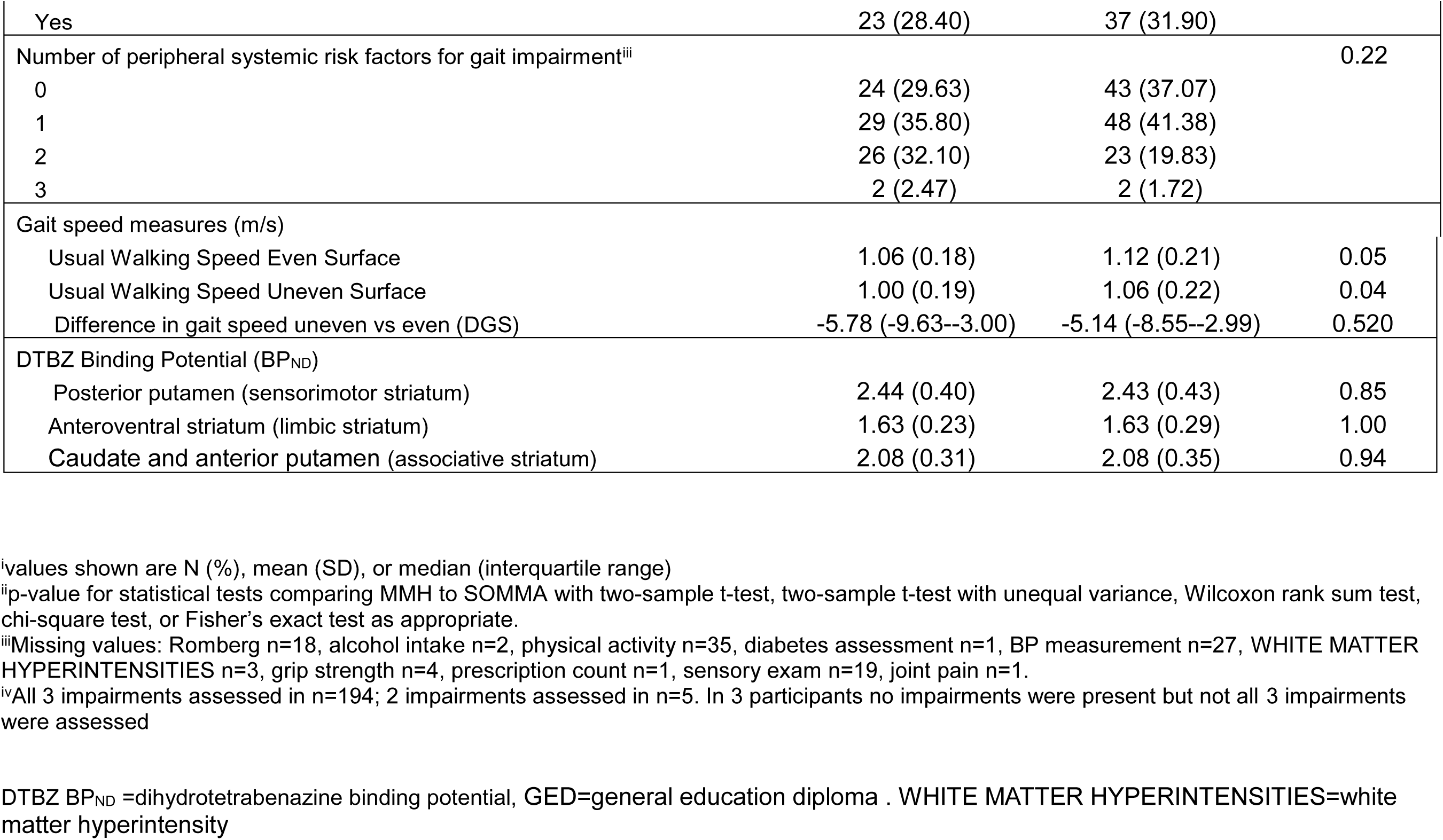
Comparison of characteristics of subsamples from each parent cohort.

| Variable<sup>i</sup> | MYHAT<br>N=81 | SOMMA<br>N=116 | p-value<sup>ii</sup> |
| --- | --- | --- | --- |
| Age (years) | 72.51 (4.50) | 76.60 (3.73) | <0.001 |
| Sex |  |  | 0.25 |
| Male | 27 (33.33) | 48 (41.38) |  |
| Female | 54 (66.67) | 68 (58.62) |  |
| Self-Identified Race |  |  | 0.88 |
| White | 74 (91.36) | 105 (90.52) |  |
| Black | 6 (7.41) | 9 (7.76) |  |
| Asian | 0 (0.00) | 1 (0.86) |  |
| Native American/Alaskan Native | 1 (1.23) | 0 (0.00) |  |
| Unknown | 0 (0.00) | 1 (0.86) |  |
| Education |  |  | <0.001 |
| Highschool/General Education Development | 25 (30.86) | 7 (6.03) |  |
| More than Highschool | 56 (69.14) | 108 (93.10) |  |
| Other | 0 (0.00) | 1 (0.86) |  |
| Smoking status |  |  | 0.034 |
| Never | 38 (46.91) | 67 (57.76) |  |
| Past | 34 (41.98) | 46 (39.66) |  |
| Current | 9 (11.11) | 3 (2.59) |  |
| Body mass index (kg/m<sup>2</sup>) | 30.91 (5.97) | 26.85 (4.37) | <0.001 |
| Alcohol Intake Past Year |  |  | 0.40 |
| No | 17 (21.25) | 31 (26.96) |  |
| Yes | 63 (78.75) | 84 (73.04) |  |
| Physical Activity: Hours Walked/Week | 0.00 (0.00-2.00) | 1.75 (0.00-3.75) | <0.001 |
| Depression Symptoms | 0.00 (0.00-1.00) | 0.00 (0.00-1.00) | 0.32 |
| Diabetes |  |  | 0.011 |
| No | 59 (73.75) | 102 (87.93) |  |
| Yes | 21 (26.25) | 14 (12.07) |  |
| Systolic Blood Pressure | 133.04 (17.16) | 130.66 (15.55) | 0.37 |
| Diastolic Blood Pressure | 75.17 (8.41) | 70.20 (8.59) | <0.001 |
| Pulse | 64.15 (8.63) | 66.46 (10.24) | 0.16 |
| Medication Count | 3.00 (2.00-5.00) | 4.00 (2.00-7.00) | 0.29 |
| Treated with antidepressant | 18 (22.22) | 19 (16.38) | 0.30 |
| Number of comorbidities |  |  | 0.28 |
| 0 | 37 (46.25) | 60 (51.72) |  |
| 1 | 31 (38.75) | 47 (40.52) |  |
| 2 or 3 | 12 (15.00) | 9 (7.76) |  |
| Positive Romberg sign<sup>ii</sup> |  |  |  |
| No | 78 (100.00) | 116 (100.00) |  |
| Yes |  |  |  |
| Vibratory sense reduced in toes<sup>ii</sup> |  |  | 0.37 |
| No | 34 (43.59) | 43 (37.07) |  |
| Yes | 44 (56.41) | 73 (62.93) |  |
| Pinprick sensation reduced in toes<sup>ii</sup> |  |  | 0.16 |
| No | 75 (94.94) | 114 (99.13) |  |
| Yes | 4 (5.06) | 1 (0.87) |  |
| Light touch sensation reduced<sup>ii</sup> |  |  | 0.065 |
| No | 76 (96.20) | 116 (100.00) |  |
| Yes | 3 (3.80) | 0 (0.00) |  |
| White matter hyperintensities (Normalized, mm<sup>3</sup><sup>iii</sup>) | 0.006 (0.003-0.009) | 0.008 (0.004-0.018) | 0.027 |
| Low Grip Strength<sup>ii</sup> |  |  | 0.96 |
| No | 54 (70.13) | 81 (69.83) |  |
| Yes | 23 (29.87) | 35 (30.17) |  |
| Obese |  |  | <0.001 |
| No | 40 (49.38) | 88 (75.86) |  |
| Yes | 41 (50.62) | 28 (24.14) |  |
| Pain for ≥1 month in hip or leg<sup>ii</sup> |  |  | 0.60 |
| No | 58 (71.60) | 79 (68.10) |  |
| Yes | 23 (28.40) | 37 (31.90) |  |
| Number of peripheral systemic risk factors for gait impairment<sup>iii</sup> |  |  | 0.22 |
| 0 | 24 (29.63) | 43 (37.07) |  |
| 1 | 29 (35.80) | 48 (41.38) |  |
| 2 | 26 (32.10) | 23 (19.83) |  |
| 3 | 2 (2.47) | 2 (1.72) |  |
| Gait speed measures (m/s) |  |  |  |
| Usual Walking Speed Even Surface | 1.06 (0.18) | 1.12 (0.21) | 0.05 |
| Usual Walking Speed Uneven Surface | 1.00 (0.19) | 1.06 (0.22) | 0.04 |
| Difference in gait speed uneven vs even (DGS) | -5.78 (-9.63--3.00) | -5.14 (-8.55--2.99) | 0.520 |
| DTBZ Binding Potential (BP<sub>ND</sub>) |  |  |  |
| Posterior putamen (sensorimotor striatum) | 2.44 (0.40) | 2.43 (0.43) | 0.85 |
| Anteroventral striatum (limbic striatum) | 1.63 (0.23) | 1.63 (0.29) | 1.00 |
| Caudate and anterior putamen (associative striatum) | 2.08 (0.31) | 2.08 (0.35) | 0.94 |

<sup>i</sup>values shown are N (%), mean (SD), or median (interquartile range)
<sup>ii</sup>p-value for statistical tests comparing MMH to SOMMA with two-sample t-test, two-sample t-test with unequal variance, Wilcoxon rank sum test, chi-square test, or Fisher's exact test as appropriate.
<sup>iii</sup>Missing values: Romberg n=18, alcohol intake n=2, physical activity n=35, diabetes assessment n=1, BP measurement n=27, WHITE MATTER HYPERINTENSITIES n=3, grip strength n=4, prescription count n=1, sensory exam n=19, joint pain n=1.
<sup>iv</sup>All 3 impairments assessed in n=194; 2 impairments assessed in n=5. In 3 participants no impairments were present but not all 3 impairments were assessed
DTBZ BP<sub>ND</sub> =dihydrotetrabenazine binding potential, GED=general education diploma . WHITE MATTER HYPERINTENSITIES=white matter hyperintensity

**Supplementary Table 2.** Relationship between usual gait speed on even and uneven surface and DTBZ binding in those with vs without any peripheral systemic risk factor for gait slowing, adjusted for age and sex.

|  | Total sample |  |  | p-value for interaction term<sup>ii</sup> |
| --- | --- | --- | --- | --- |
|  | sβ | SE | p-value<sup>i</sup> |  |
| Outcome: usual gait speed on even surface |  |  |  |  |
| Predictor: [<sup>11</sup>C]DTBZ BP<sub>ND</sub> in striatal region |  |  |  |  |
| Posterior putamen (sensorimotor striatum) | 0.116 | 0.073 | 0.11 | 0.44 |
| Anteroventral striatum (limbic striatum) | 0.016 | 0.076 | 0.84 | 0.81 |
| Caudate and anterior putamen (associative striatum) | 0.033 | 0.073 | 0.65 | 0.72 |
| Outcome: usual gait speed on uneven surface |  |  |  |  |
| Predictor: [<sup>11</sup>C]DTBZ BP<sub>ND</sub> in striatal region |  |  |  |  |
| Posterior putamen (sensorimotor striatum) | 0.026 | 0.015 | 0.09 | 0.89 |
| Anteroventral striatum (limbic striatum) | 0.004 | 0.016 | 0.82 | 0.62 |
| Caudate and anterior putamen (associative striatum) | 0.011 | 0.015 | 0.48 | 0.89 |

<sup>i</sup> Linear regression model consisting of: difference in specified gait speed measure (outcome), [<sup>11</sup>C]DTBZ BP<sub>ND</sub> in specified striatal region, age, and sex
<sup>ii</sup> Linear regression model consisting of: difference in gait speed measure (outcome), [<sup>11</sup>C]DTBZ BP<sub>ND</sub> in specified striatal region, age, and sex, presence of any risk factor, and interaction term between risk factor present (yes/no) and [<sup>11</sup>C]DTBZ BP<sub>ND</sub> in specified striatal region
DTBZ BP<sub>ND</sub> =dihydrotetrabenazine binding potential. PSRF=peripheral systemic risk factors. sβ=standardized beta coefficient. SE=standard error

